# Enhanced Myocardial Tissue Visualization: A Comparative Cardiovascular Magnetic Resonance Study of Gradient-Spin Echo-STIR and Conventional STIR Imaging

**DOI:** 10.1101/2023.10.24.23297504

**Authors:** Sadegh Dehghani, Shapoor Shirani, Elahe Jazayeri gharehbagh

## Abstract

**Purpose:** To assess the performance of gradient-spin echo (GraSE) based STIR (GraSE-STIR) sequence in CMR imaging compared to turbo spin echo based conventional STIR for myocardial visualization.

**Methods:** In a prospective study, we examined forty-four normal volunteers and seventeen patients referred for CMR imaging using a conventional STIR and GraSE-STIR techniques. Signal-to-noise ratio (SNR), contrast-to-noise ratio (CNR), image quality, T_2_ signal intensity (SI) ratio, specific absorption rate (SAR), and image acquisition time were compared between both sequences.

**Results:** GraSE-STIR showed significant improvements in image quality (4.15 ± 0.8 vs. 3.34 ± 0.9, P = 0.024) and cardiac motion artifact reduction (7 vs. 18 out of 53, p = 0.038) compared to conventional STIR. Furthermore, the acquisition time (27.17 ± 3.53 vs. 36.9 ± 4.08 seconds, p = 0.041) and the local torso SAR (< % 13 vs. < % 17, p = 0.047) were significantly lower for GraSE-STIR compared to conventional STIR in short axis plan. However, no significant differences were shown in T_2_ SI ratio (p = 0.141), SNR (p = 0.093), CNR (P = 0.068), and SAR (p = 0.071) between these two sequences.

**Conclusions:** GraSE-STIR offers notable advantages over conventional STIR sequence, with improved image quality, reduced motion artifacts, and shorter acquisition times. These findings highlight the potential of GraSE-STIR as a valuable technique for routine clinical CMR imaging.

## 1. Introduction

Advances in cardiovascular magnetic resonance (CMR), using short tau inversion recovery (STIR), have allowed for the visualization of regional and global myocardial edema in various heart diseases.^1, 2^ The conventional STIR, as a T_2_-weighted turbo spin echo (T_2_-w TSE) or edema imaging technique, uses a third inversion pulse following a double inversion recovery (DIR) black blood preparation to additionally null the fat signal.^3, 4^ Therefore, by using triple inversion recovery (TIR), conventional STIR black-blood technique aids to diagnose acute myocardial lesions more precisely.^5, 6^ In this technique, given the non-spectrally selective third inversion pulse, signal generated by water protons in the myocardium is also inverted, resulting in improved contrast-to-noise ratio (CNR). As a result, it enhances the differentiation between edematous tissue and normal myocardium.^7, 8^ However, the conventional STIR is TSE based sequence with TSE factor = 25-35. High TSE factors present several challenges in this technique, including increased sensitivity to slow-flow artifacts around the endocardial borders, resulting in elevated signal intensity (SI),^9^, ^10^ and cardiac motion artifacts.^11^ Moreover, a high TSE factors is subject to high specific absorption rate (SAR).^12^

Gradient-spin echo (GraSE) technique may offer an alternative to conventional TSE imaging. The GraSE technique is a fast MRI method that combines elements of the TSE and echo planar imaging (EPI) techniques. It involves generating a series of spin echoes using multiple 180° refocusing radiofrequency (RF) pulses, where gradient echoes are sampled before and after each spin echo with an EPI readout.^13^ By employing EPI as a gradient-echo readouts (GRE) technique, leading to increased speed in TSE sequences, the energy deposition in the patient can be reduced.^14^ Furthermore, the acquisition time will be reduced, resulting in lower cardiac motion artifacts, particularly for patients with irregular breathing patterns.^15^

The goal of our study was therefore to systematically compare GraSE-STIR and conventional STIR techniques in a cohort of normal volunteers and patients with cardiac edema disease aiming to analyze the impact of GraSE technique on myocardial SI, image quality, safety parameters, and acquisition time.

## 2. Methods

The local ethic committee approved this prospective study and written informed consent was obtained from all volunteers prior to CMR imaging. All scans were performed using a 1.5 tesla (T) MRI system (Ingenia, Philips Medical Systems, Best, The Netherlands) with a maximum gradient strength of 45 mT/m and a maximum slew rate of 200 mT/m/ms. A 32 channel torso coil with digital interface was used for signal reception.

### 2.1. Study cohort

We assessed forty-four normal volunteers (mean age of 44.8 ± 19.6 years, 20 females and 24 males) with no cardiac disease history. Furthermore, seventeen patients with cardiac edema disease were included to this study (mean age of 41.7 ± 14.8 years, 7 female patients and 10 male). Participants were scanned with both conventional STIR and GraSE-STIR protocols for the purposes of comparing T_2_ SI ratio, SNR, CNR, image quality, and artifacts. Of these fifty-three volunteers, CNR was only calculated in eleven patients who had focally increased SI in edema imaging techniques. Exclusion criteria included magnetic resonance incompatible implants, severe claustrophobia, and general contraindications to MRI, clinical instability, and sever artifacts in desired images, leading to bias in qualitative and quantitative measurement. Characteristics of patients were summarized in Table 1.

**Table 1.** Characteristics of patients.

| **Parameters** | **Results** |
| --- | --- |
| **Number of normal volunteers (females/males)** | 44 (20/24) |
| **Age (years)** | 44.8 ± 19.6 |
| **Number of patients (females/males)** | 17 (7/10) |
| **Age (years)** | 41.7 ± 14.8 |
| **Heart rate (min<sup>-1</sup>)** | 66.13 ± 18.57 |
| **Height [cm]** | 160.73 ± 16.47 |
| **Weight [kg]** | 60.54 ± 19.21 |
| **LV<sup>a</sup> ejection fraction [%]** | 53 ± 13.5 |
| **LV end-diastolic volume / BSA<sup>b</sup> [ml/m<sup>2</sup>]** | 68.7 ± 16.48 |
| **LV end-systolic volume / BSA [ml/m<sup>2</sup>]** | 29.57 ± 8.4 |

<sup>a</sup>LV: left ventricular,
<sup>b</sup>BSA: body surface area

### 2.2. MRI protocol

The imaging protocol included cine, using balanced steady-state free precession (bSSFP), conventional STIR, and GraSE-STIR sequences at standardized apical, mid-cavity, and basal short-axis (SAX) levels,^16^ covering 16 segments of the 17-segment model as recommended by American Heart Association (AHA),^17^ two chamber (2CH), three chamber (3CH), as well as four chamber (4CH) plans. The late gadolinium enhancement (LGE) imaging was also done for patients with cardiac disease. All images were acquired, using following parameters: For bSSFP and LGE, acquired pixel size = 1.8 × 2.3 mm, reconstructed pixel slice = 1.5 × 1.5 mm, slice thickness = 8 mm during end-expiration breathhold period. For cine imaging, bSSFP pulse sequence was used (TR = 3.5 ms, TE = 1.4 ms, FA = 55°, and temporal resolution ≤ 45 ms). For conventional STIR, we used a TIR black-blood turbo-spin echo pulse sequence with TSE factor = 25 and TSE_es_ = 5.4 ms. For GraSE-STIR, we used a TIR black-blood GraSE pulse sequence with TSE factor = 18, EPI factor = 3, and TSE echo spacing (TSE_es_) = 7.4 ms. Other parameters, including TR = 2 heartbeat, TE = 80 ms, TI = 165 ms, slice thickness = 10 mm, field of view (FOV) = 270×270 mm, matrix size = 124×122, acceleration factor = 1.4, shot duration = 134 ms, volume shimming, and bandwidth = 204 Hz/pixel were the same for both sequences. The pulse sequence diagrams for conventional STIR and GraSE-STIR sequences were illustrated in figure 1. In case of patients with cardiac disease, eight minutes after intravenous application of a standard MRI contrast agent (DOTAREM, 0.4 mmol/kg), look locker sequence was performed with increasing TI to achive the nulling of normal myocardium. After that, definitive LGE images were acquired with an IR-prepared gradient echo technique (TR = 4.5 ms, TE = 1.8 ms, and FA = 15), using the TI that allowed for the best nulling of normal myocardium.

**Fig. 1.**
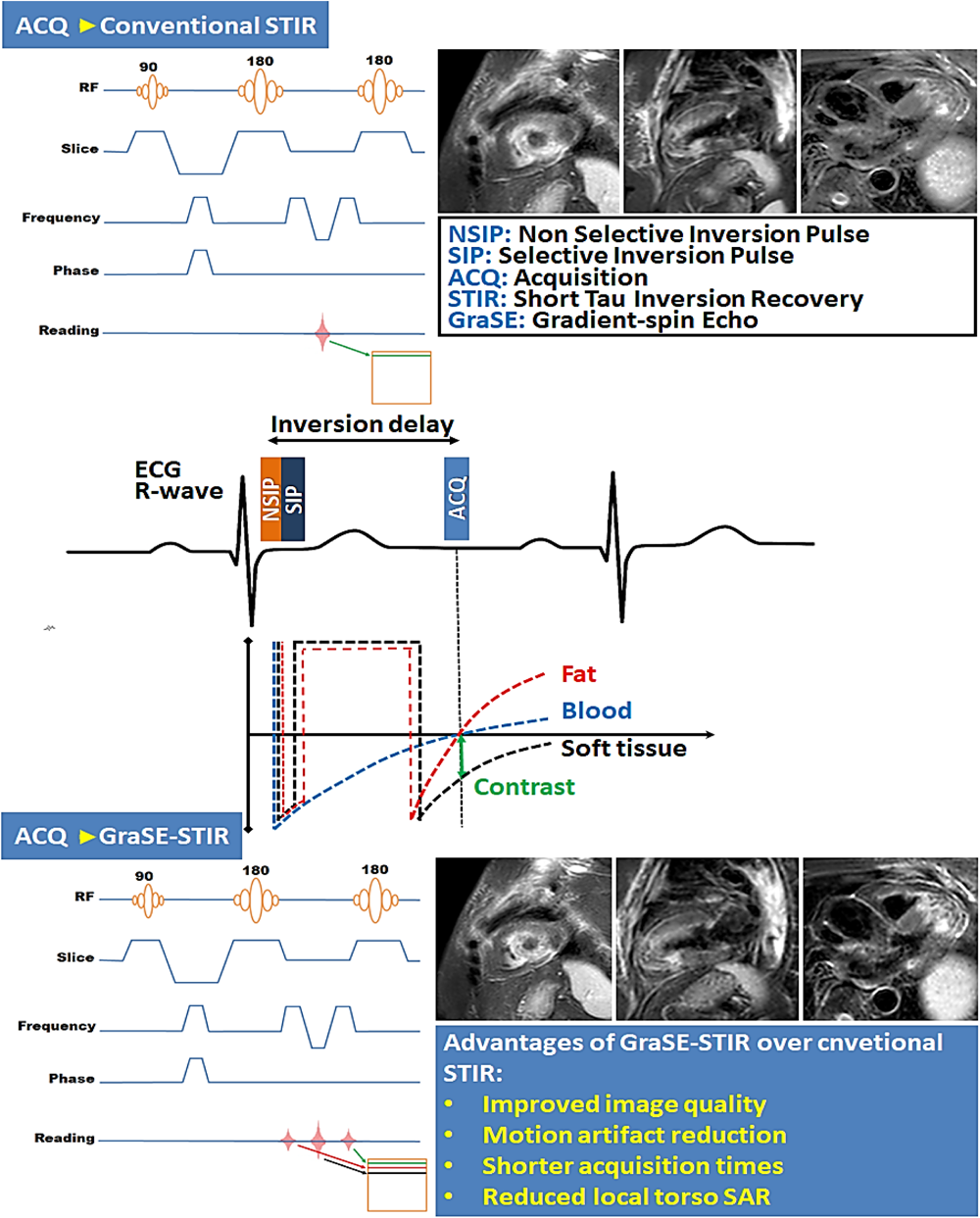
The pulse sequence diagrams and associated images for conventional STIR and gradient-spin echo (GraSE) based STIR (GraSE-STIR) sequences are schematically depicted

### 2.3. Image analysis

#### 2.3.1. T_2_ SI ratio measurement

To assess the presence of edema, the T_2_ SI ratio of left ventricular (LV) myocardium to skeletal muscle was calculated for all T_2_w images with Philips extended MR work space (version 2.6.3.4; Philips Medical Systems, Best, The Netherlands). According to previous study, a T_2_ SI ratio equal to or above a threshold of two was considered to be positive for the presence of edema.^2^ Calculation of the T_2_ SI ratio involved manually tracing region of interest (ROI) contours in different plans of the conventional STIR and GraSE-STIR images. In every plan, five ROIs were traced on different parts of LV myocardium and five ROIs on skeletal muscles and then the mean SI for LV myocardium and skeletal muscle was calculated. In the presence of edema, we measured the T_2_ SI ratio in a single myocardial slice per patient, specifically in the slice with the most extensive regional edema. To avoid slow-flow artifacts, ROIs were traced by carefully omitting the inner endocardial border. In all T_2_w images, skeletal muscle was identified as cross-referenced by cine images in the same slice position. The mean SI of the LV myocardium was divided by the mean SI of skeletal muscle in the same slice resulting in a T_2_ SI ratio.^18^

#### 2.3.2. CNR measurement

Relative CNR was calculated in patients who had evidence of regional SI increase in T_2_-w images, representing edema. The mean SI of the edematous region was subtracted from mean SI of remote myocardium, and divided by the mean standard deviation (SD) of background noise.^19^ The mean SD of background noise was obtained by tracing a ROI in air 2–3 cm anterior to the chest wall.^20^ CNR was only performed in one myocardial slice per patient, where the greatest extent of regional edema was present.

In order to compare slow-flow artifact suppression between conventional STIR and GraSE-STIR, CNR was calculated between myocardium and blood. The LV myocardial mean SI was subtracted from the mean SI of blood within the LV myocardium and divided by mean SI of blood.^19^

#### 2.3.3. SNR measurement

Relative SNR was calculated for conventional STIR and GraSE-STIR images. For calculating SNR, the mean SI of LV myocardium was divided by the mean SD of the background noise for each slice.^20^

### 2.4. Image quality analysis

Two experienced CMR observers, each with over 5 and 8 years of experience, independently and blindly assessed T_2_w images from patients. Their analysis involved the qualitative evaluation of myocardial border clarity, as well as the identification of artifacts, including high SI at the subendocardial border of the LV myocardium, posterolateral signal loss, and cardiac motion that might affect the LV myocardium assessment and interpretation. Image quality was scored using the following 5-point rating scale:^21^ 1 represented severe image distortion from artifacts that significantly limited interpretation, 2 indicated significant impairment due to artifacts, 3 denoted moderate artifact impairment, 4 reflected mild artifact impairment, and 5 represented images without impairment from artifacts. In cases where a consensus could not be reached between the two radiologists, a third radiologist with ten years of experience in cardiac MRI was consulted to resolve the issue. Additionally, above mentioned artifacts were assessed separately, identifying the presence of any artifact in each image, and the number of patients exhibiting each type of artifact was calculated. Finally, the scan time required for both conventional STIR and GraSE-STIR sequences was evaluated.

### 2.5. Inter-and intra-rater repeatability

In this study, the intra-rater repeatability was calculated from two independent evaluations of all subjects by a single observer, separated by one month and it was expressed as an intra class correlation coefficient, (ICC). Furthermore, the inter-rater reliability coefficient (IRR) was used to show the agreement among raters as a basis for our calculation.^22^

### 2.6. Statistical analysis

Data analysis was done using Origin pro 2022 version. Descriptive statistics, such as means, SD, and confidence intervals, were calculated. Paired t-tests was used to assess differences between the sequences. The Pearson rank correlation analyses were conducted to evaluate the associations between image qualities measured by observers. Kappa values were those between 0 and 0.40, poor agreement; those between 0.41 and 0.75, good agreement; and those between 0.76 and 1.00, excellent agreement.^23^

## 3. Results

T_2_-w imaging was done, using conventional STIR and GraSE-STIR sequences. Eight volunteers (4 normal volunteers and 4 patients) were excluded from analysis due to non-eligible visual artefacts in both sequences. The remaining forty normal volunteers and thirteen patients formed the study group for measuring of GraSE-STIR performance.

### 3.1. Image quality analysis

Significant improvement in image quality was observed with GraSE-STIR compared to conventional STIR sequence (4.15 ± 0.8 vs. 3.34 ± 0.9, P = 0.024). The assessment of intra-and inter-reader agreement for both sequences revealed very good agreement between repeated readings of the same reader (ICC > 0.74) and between the measurements of different readers (IRR > 0.71). Figure 2 shows high-quality images of conventional STIR and GraSE-STIR sequences from normal volunteer.

**Fig. 2.**
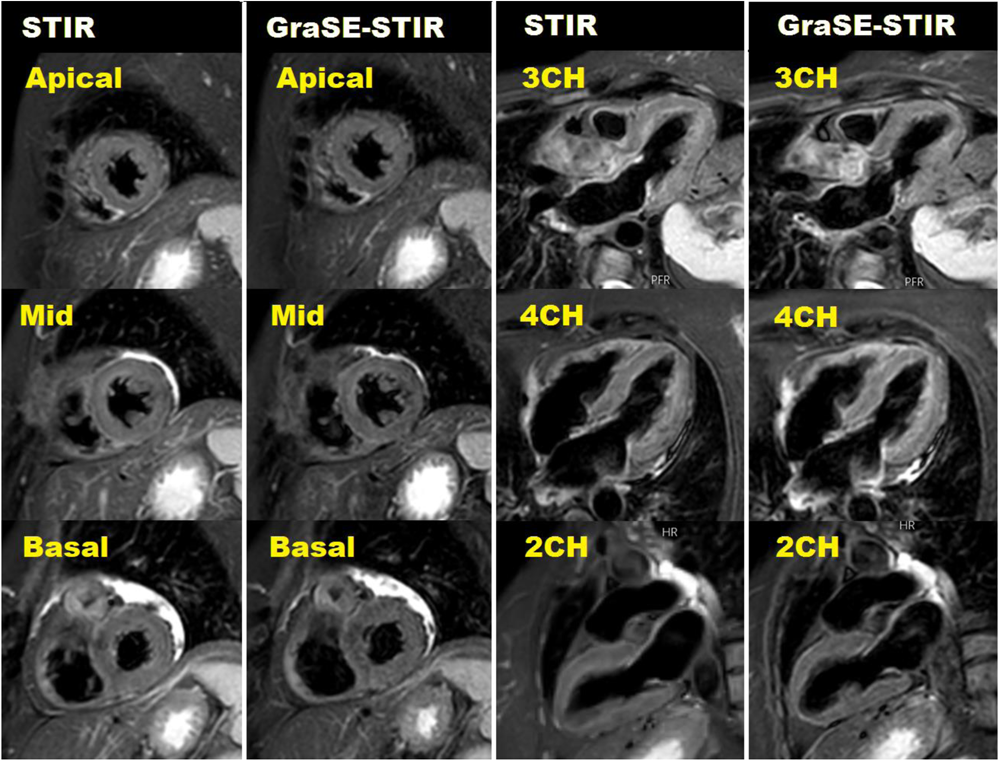
GraSE-STIR and conventional STIR images of normal male volunteer. Image quality analysis represented high-quality images (scale of 5 for both techniques) for both sequences.

### 3.2. Artifacts measurement

High SI artifacts due to slow-flow of blood next to the endocardium in LV myocardium was observed in 26 and 24 out of 53 volunteers in conventional STIR and GraSE-STIR, respectively, with no significant differences (p = 0.078). As shown in figure 3, cardiac motion artifact was significantly higher in conventional STIR compared to GraSE-STIR sequence (18 vs. 7 out of 53, p = 0.038). Posterolateral loss of signal in conventional STIR (3 out of 53) and GraSE-STIR (5 out of 53) sequences did not show any statistical differences (p = 0.082).

**Fig. 3.**
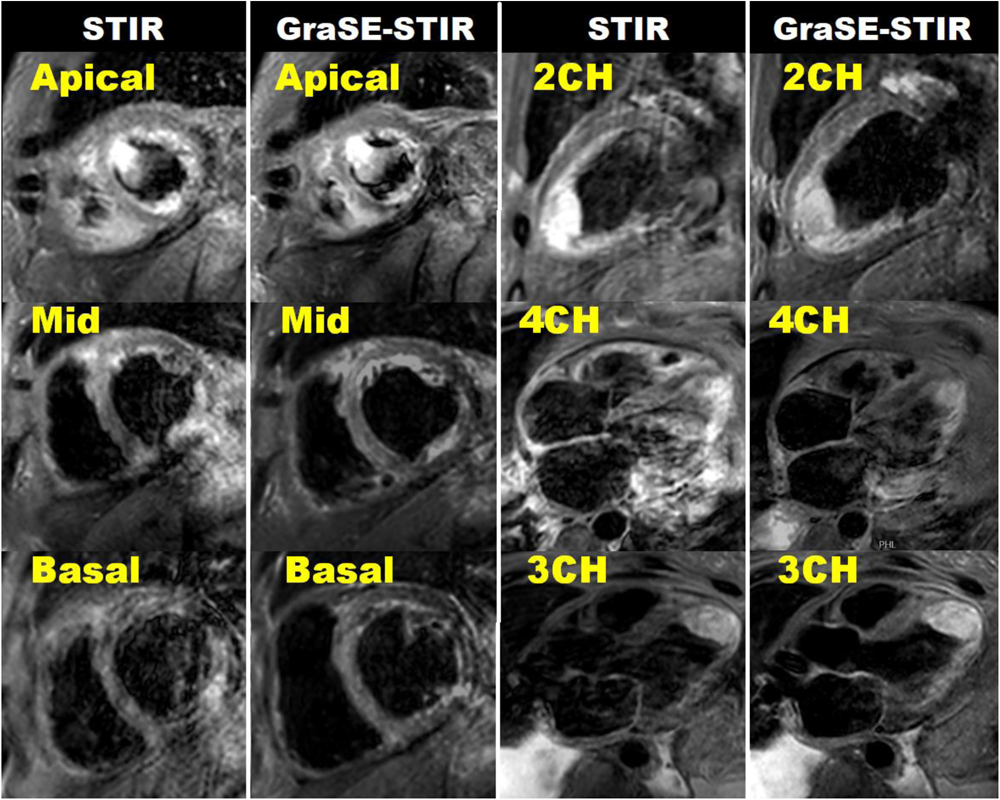
GraSE-STIR and conventional STIR images of patient with severely enlarged size and reduced systolic function. Large semi mobile clot with edema can be seen in apical section. The image quality of GraSE-STIR (scale of 4 out of 5) is significantly higher than conventional STIR sequence (scale of 3 out of 5). Motion artifact is clearly seen in 2CH and 4Ch of conventional STIR images. High SI artifacts in LV myocardium is clearly seen in SAX, 2CH, and 4CH plans of both sequences with no significant differences.

### 3.3. Quantitative analysis

#### 3.3.1. T_2_ SI ratio

The T_2_ SI ratio between myocardial edema and musculoskeletal muscle was achieved 2.5 ± 1.2 for conventional STIR and 2.4 ± 0.8 for GraSE-STIR sequence. No significant differences were observed between these two sequences, and good correlations were obtained between both techniques (p = 0.71, ICC = 0.64, IRR = 0.56). T_2_ SI ratios were also measured between normal myocardium and musculoskeletal muscle for both sequences in all planes. T_2_ SI ratios and Pearson correlations were summerized in Table 2. No significant differences were found between both sequences (p = 0.141). Additionally, a good correlation was achieved between sequences (ICC = 0.70 – 0.80, IRR = 0.62 – 0.72).

**Table 2.** Quantitative analysis of T_2_ signal intensity (SI) ratio of normal myocardial tissue.

| **T<sub>2</sub> signal**<br>**intensity ratio** | **Conventional**<br>**STIR** | **GraSE-STIR** | **Intra-rater**<br>**correlation (ICC)** | **Intr-rater**<br>**correlation**<br>**(IRR)** |
| --- | --- | --- | --- | --- |
| **Basal** | 1.79 ± 0.2 | 1.77 ± 0.17 | 0.71 | 0.62 |
| **Mid-ventricle** | 1.76 ± 0.15 | 1.75± 0.21 | 0.77 | 0.65 |
| **Basal** | 1.78 ± 0.18 | 1.74± 0.23 | 0.80 | 0.72 |
| **2CH** | 1.77 ± 0.16 | 1.79± 0.16 | 0.75 | 0.68 |
| **3CH** | 1.81 ± 0.13 | 1.77± 0.14 | 0.7 | 0.62 |
| **4CH** | 1.81± 0.14 | 1.79± 0.2 | 0.72 | 0.66 |
| **Global** | 1.78 ± 0.17 | 1.77± 0.19 | 0.7 | 0.65 |

#### 3.3.2. CNR measurement

In patients with regional myocardial edema, CNR between edema and remote normal myocardium demonstrated no significant difference between GraSE-STIR (116 ± 14.51) and conventional STIR (108 ± 20) sequences with a p-value of 0.068. Furthermore, CNR between myocardium and blood showed no significant differences between two sequences (8.36 ± 5.12 vs. 9.5 ± 4.21, P = 0.052). Figure 4 shows CMR images of patient with myocardial edema.

**Fig. 4.**
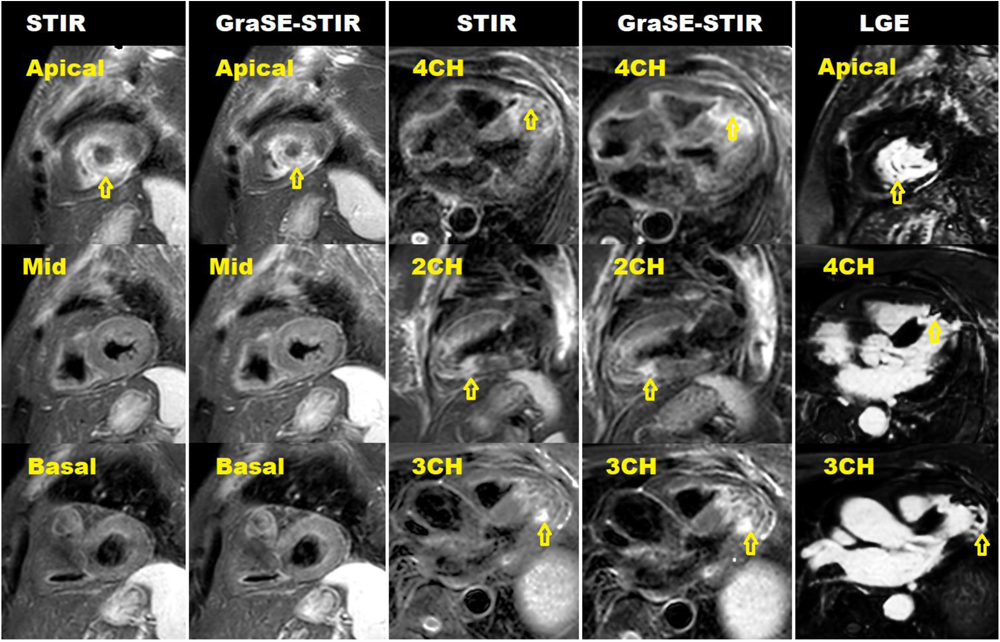
The comparison of myocardial edema imaging techniques. GraSE-STIR, conventional STIR, and late gadolinium enhancement (LGE) of myocardial edema in patient with normal left ventricle (LV) size, reduced systolic dysfunction and apical segments aneurysm formation with layered apical clot. Regional edema is defined by areas of increased SI (arrows) in all sequences.

#### 3.3.3. SNR measurement

SNR of conventional STIR sequence for all plans was calculated as follow: Apical, 179 ± 13.61; mid-ventricle, 180 ± 12.5; basal, 182 ± 13.17; 2CH, 177.55 ± 9.78; 3CH, 175.87 ± 12.23; and 4CH, 181.38 ± 10.15. No significant differences were shown a cross all plans (p = 0.116). SNR of GraSE-STIR sequence was calculated as follow: Apical, 176 ± 9.23; mid-ventricle, 180.24 ± 8.17; basal, 174 ± 9.18; 2CH, 179.78 ± 11.48; 3CH, 180 ± 9.97; and 4CH, 177.31 ± 9.69. No significant differences were shown a cross all plans (p = 0.106). Furthermore, no significant differences were shown between two sequences (p = 0.093, fig. 5a).

**Fig. 5.**
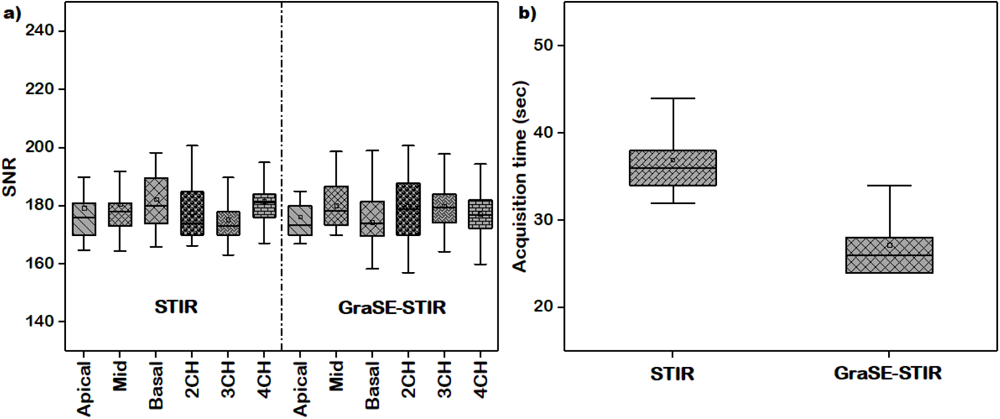
Quantitative analysis of Conventional STIR and GraSE-STIR in CMR imaging. a) Signal-to-noise ratio (SNR) of conventional STIR and GraSE-STIR. No significant differences were found among different planes for conventional STIR (p = 0.116) and GraSE-STIR (p = 0.106) as well as between both sequences (p = 0.093). b) Time taken for conventional STIR was significantly higher than GraSE-STIR in short axis (SAX) view (p = 0.041).

#### 3.3.4. Acquisition time measurement

The acquisition time for every plan of 2ch, 3ch, and 4ch was 6-8 seconds for GraSE-STIR sequence and 8-12 seconds for conventional STIR without any significant differences (p = 0.061). However, as demonstrated in figure 5b, the mean duration of time taken for GraSE-STIR and conventional STIR sequences was 27.17 ± 3.53 and 36.9 ± 4.08 seconds, respectively, in SAX plans with statistically significant difference (p = 0.041).

### 3.4. SAR measurement

In conventional STIR imaging, SAX view, SAR was measured to be less than 0.4 W/kg, and the local torso SAR was successfully limited to less than 17%. However, in GraSE-STIR imaging, SAR was achieved less than 0.3 W/kg and local torso SAR was limited to less than 13%. Our statistical analysis revealed no significant differences in SAR (p = 0.071). While local torso SAR was statistically lower in GraSE-STIR sequence (p = 0.047). No variation was found for SAR (< 0.3 W/kg) and local torso SAR (< 10%) in 2CH, 3CH, and 4CH for both STIR and GraSE-STIR sequences (p = 1.00).

## 4. Discussion

The utilization of CMR has revolutionized the understanding and diagnosis of myocardial pathologies, offering non-invasive insights into tissue characteristics. One key aspect in CMR imaging is the accurate visualization of myocardial edema using STIR technique, which aids in differentiating acute from chronic myocardial lesions. However, the conventional STIR black-blood technique, though widely employed, has been associated with varying image quality, raising concerns about its diagnostic reliability in clinical practice.^5, 6^

### 4.1. Image acquisition time

Although, conventional STIR plays an important role for edema imaging, the long acquisition time of this sequence, as it is based on TSE technique, can result in cardiac motion artifact especially in tachyarrhythmia patients, leading to image quality reduction.^24^ While GraSE sequence, combining advantages of EPI and TSE techniques, has been shown to allows for robust, reliable, and fast myocardial T_2_ mapping in CMR imaging, and quantitative tissue characterization imaging.^20, 21^ GRASE imaging was also shown to reduce scan time for brain imaging and have a potential for clinical use in patients with brain disorders.^25, 26^ In this study, GraSE-STIR demonstrated a significantly shorter acquisition time in the SAX view compared to conventional STIR, leading to significantly higher image quality.

### 4.2. Quantitative analysis

The T_2_ SI ratio for conventional STIR and GraSE-STIR sequences in all plans for participants without edema remained within the normal range, with no significant differences. Furthermore, T_2_ SI ratio between myocardial edema and musculoskeletal showed no significant differences between both sequences that was aligned with previous studies.^27, 28^ This provides the assurance that GraSE-STIR maintains image quality standards. In our study, the same SNR observed with GraSE-STIR, compared to conventional STIR, across different myocardial segments highlights its efficacy in capturing detailed tissue information like conventional STIR. Furthermore, the same CNR observed in patients with regional myocardial edema further emphasizes the diagnostic potential of GraSE-STIR, similar to conventional STIR. Bagnato, F., et al.^29^ showed that GRASE sequences with EPI factor/TE = 3/50 and 3/75 ms were comparable to the T_2_-w TSE technique for measuring the CNR between white matter lesions and normal-appearing white matter as well as detection of white matter lesions. Giri S., et al^30^ and Bönner F., et al^31^ also revealed only minor regional variations in T_2_ relaxation times in healthy volunteers when using the GraSE technique compared to other T_2_ mapping techniques. These findings emphasize the reliability and reproducibility of the GraSE technique for edema imaging.

### 4.3. Image quality analysis

Image quality indeed plays an important role in achieving accurate diagnoses.^32^ Therefore, it’s important to highlight the significant improvement in image quality and motion artifact reduction achieved with GraSE-STIR when compared to conventional STIR. In this study, the excellent agreement between independent observers reinforced the reliability of the GraSE-STIR technique in producing consistent image quality. Additionally, GraSE-STIR maintained comparable values for SNR, CNR, slow-flow artifact, and posterolateral signal loss when compared to conventional STIR. This suggests that GraSE-STIR can provide detailed tissue information compared to conventional STIR.

### 4.4. SAR and local torso SAR measurements

A crucial aspect of MRI studies is SAR, reflecting energy deposition in the patient’s tissue during scans. SAR reduction is especially beneficial in scenarios involving repeated or extended imaging, minimizing tissue heating and patient discomfort.^33^ Our findings demonstrate that the GraSE-STIR sequence not only offered enhanced image quality but also exhibited a reduction in local torso SAR in SAX view compared to the conventional STIR sequence. Although, no significant differences were observed between GraSE-STIR and conventional STIR in terms of SAR, this difference can be significant in higher magnetic fields. It is important to note that at higher magnetic fields (e.g., 7 T), obtaining T_2_-w TSE images in vivo is not possible due to the high SAR.^29^ Therefore, techniques reducing the SAR without compromising image quality could be a potential alternative for TSE based techniques. Okanovic M, et al.^34^ and Chu ML, et al.^35^ also confirmed our work. They indicated high-quality, artifact-free images with a significant SAR reduction in GRASE in magnetic fields exceeding 3 T compared to TSE sequence.

### 4.5. Limitations

Firstly, it was conducted at a single center utilizing a specific MRI scanner, potentially constraining the broader applicability of our edema imaging methodology. Multicenter studies are needed to validate the reproducibility and transferability of our findings. Secondly, the sample size in our study was small, primarily due to the long acquisition time required for CMR scans. This limited our ability to incorporate a larger cohort of volunteers. Thirdly, most of volunteers were normal with approximately normal heart rate. While, this technique can be evaluated better in tachyarrhythmia patients with severe cardiac motion artifact. Fourthly, these two techniques were compared in 1.5 T MRI scanner, while at higher magnetic fields, GraSE-STIR and conventional STIR may be more comparable in terms of SAR. These limitations highlight the need for future research to address these gaps and provide a more comprehensive understanding of edema imaging in different populations.

## 5. Conclusion

The GraSE-STIR approach not only offered enhanced image quality and motion artifact reduction but also reduced local torso SAR compared to conventional STIR imaging. Our findings highlight the safety and feasibility of incorporating GraSE-STIR into clinical practice to enhance patient comfort during scanning. By addressing both diagnostic and safety aspects, GraSE-STIR holds significant potential as a valuable tool for myocardial tissue characterization in CMR imaging.

## Disclosures

None

## Data Availability

All data referred to in the manuscript are available upon request from the corresponding author.

## Notes

### Competing Interest Statement

The authors have declared no competing interest.

### Funding Statement

No external funding was received

### Author Declarations

This study was approved by the local ethics committee at Tehran University of Medical Sciences under the reference number IR.TUMS.SPH.REC.1402.045.

